# Integrating stakeholder perspectives in modeling routine data for therapeutic decision-making

**DOI:** 10.64898/2026.02.18.26346074

**Authors:** Michelle Pfaffenlehner, Andrea Dreßing, Dietrich Knoerzer, Markus Wagner, Peter Heuschmann, André Scherag, Harald Binder, Nadine Binder, the EVA4MII project

## Abstract

**Background:** Routinely collected health data are increasingly used to generate real-world evidence for therapeutic decision-making. Yet, stakeholders, including clinicians, pharmaceutical industry representatives, patient advocacy groups, and statisticians, prioritize different aspects of data quality, analysis, and interpretation. Without explicit consideration of these perspectives, analyses risk being fragmented, misaligned with end-user needs, or lacking transparency.

**Methods:** We developed a stakeholder-inclusive conceptual framework for modeling routine health data, informed by an interdisciplinary workshop and supported by targeted literature examples. The framework maps stakeholder priorities to methodological requirements and identifies analytical strategies that enable integration of diverse perspectives.

**Results:** Clinicians prioritize interpretability and clinical relevance; the pharmaceutical industry emphasizes regulatory compliance and real-world evidence generation; patient groups highlight transparency, inclusion of patient-reported outcomes, and privacy protection; and statisticians focus on bias control and methodological rigor. Our framework illustrates how these priorities can be explicitly incorporated into modeling strategies. Multistate models exemplify a methodological approach that operationalizes these requirements by capturing dynamic disease trajectories, integrating intermediate outcomes, and offering graphical interpretability. Beyond specific methodological choices, clinical research relies fundamentally on statistical expertise. Depending on the research goal, statisticians’ roles can range from providing statistical consultations for standard analyses to applying or adapting advanced methods for more complex analyses to developing new methods for research questions that require novel approaches due to their specific characteristics.

**Conclusions:** The stakeholder-inclusive framework provides methodological guidance for designing analyses of routine health data that are clinically meaningful, scientifically rigorous, and socially acceptable. By aligning the research question with the intended perspective from the beginning, it supports more robust and transparent evidence generation, with multistate models serving as a flexible tool to operationalize this integration.

## 1. Introduction

Routinely collected health data, including electronic health records (EHR), registries, claims data, and patient-reported outcomes, are increasingly used to complement evidence from randomized controlled trials [1]. Such real-world evidence can address gaps where trials are either difficult to carry out or sometimes infeasible, for example studies with exposures than cannot be assigned experimentally, studies in vulnerable or rare disease populations, or studies that look at long-term outcomes. In Germany, the introduction of the Health Data Use Act (GDNG) in 2024 has further expanded opportunities for analyzing routine data, supported by nationwide initiatives to standardize and harmonize electronic health record data [2–4].

Despite these opportunities and in contrast to e.g. pragmatic randomized controlled trials, the methodological use of routine data tends to depend stronger on the perspectives of multiple stakeholders. While clinicians seek clinically interpretable models that inform decision-making; the pharmaceutical industry requires robust evidence to meet regulatory standards; patient advocates emphasize transparency, inclusion of patient-reported outcomes, and data protection; and statisticians focus on validity, bias reduction, and analytical rigor. Each perspective highlights distinct priorities, yet no common framework currently exists to align these needs in the design and implementation of routine data analyses.

This article provides a stakeholder-inclusive conceptual framework for modeling routine health data. The framework explicitly maps stakeholder priorities to methodological requirements, illustrating how different perspectives can be integrated into statistical modeling. The development of this framework is depicted in Section 2. The conceptual framework is then explained in Section 3 where we first present the expectations and needs from different perspectives (see Section 3.1). In Section 3.2, we highlight multistate models (MSMs) as a particularly suitable methodological approach, given their flexibility in representing disease and treatment trajectories, handling competing risks and recurrent events, and incorporating patient-centered outcomes. In Section 4, we discuss limitations from different perspectives and limitations of this work. By combining stakeholder perspectives with methodological considerations, we aim to provide guidance for designing analyses that are both scientifically robust and responsive to the diverse needs of healthcare decision-making.

## 2. Development of the Conceptual Framework

To develop the stakeholder-inclusive conceptual framework, we employed a two-step approach. First, we gathered perspectives through an interdisciplinary workshop that brought together stakeholders from clinical medicine, biostatistics, patient advocacy, and the pharmaceutical industry, and second, to identify illustrative examples that map stakeholder priorities to analytical strategies.

### 2.1 Interdisciplinary workshop

The workshop was organized as part of the EVA4MII initiative at the annual German Association for Medical Informatics, Biometry and Epidemiology (GMDS) conference in September 2024 [5,6]. An expert panel was convened with one representative for each stakeholder perspective, including clinicians, statisticians, patient advocates, and the pharmaceutical industry (also reflected in the authorship). After short expert presentations, the discussion continued in a fishbowl format that encouraged interaction with the conference audience, which itself reflected the diversity of stakeholder perspectives typical of GMDS meetings. Notes and outputs from the session were collated and summarized thematically to identify priorities, challenges, and expectations regarding the use of routine health data for therapeutic decision-making.

### 2.2 Targeted literature review

To complement workshop insights, we conducted a targeted review of methodology and applied literature in clinical epidemiology, pharmacoepidemiology, and health services research. We focused on publications that exemplify the alignment of stakeholder priorities with analytical methods, focusing on MSMs that is a powerful tool to analyze different research topics, such as disease or treatment pathways, and is also capable to integrate patient-reported outcomes. Literature was identified through PubMed and Google Scholar searches, as well as reference lists of key methodological reviews.

### 2.3 Synthesis process

Insights from the workshop and literature review were synthesized into a conceptual matrix linking stakeholders, priorities, methodological needs, and illustrative methods. Through iterative discussion among the author team, this synthesis was refined into the stakeholder-inclusive framework presented in Section 3. The framework aims to be generalizable across routine data contexts, while MSMs are highlighted as one example of how methodological approaches can operationalize multiple stakeholder requirements.

## 3. The Stakeholder-Inclusive Conceptual Framework

Synthesizing the GMDS 2024 expert panel discussion with targeted literature examples yields a stakeholder-inclusive conceptual framework that links four core perspectives (i) clinicians, (ii) pharmaceutical industry, (iii) patient advocates, and (iv) statisticians to their priorities, methodological needs, and suitable analytic strategies illustrated by means of MSMs. Despite different emphases, common ground emerges around the demand for analyses that are clinically meaningful, transparent, reproducible, and capable of representing patient pathways and intermediate outcomes. Below, we summarize each perspective and needs (see Figure 1), and indicate afterwards how MSMs are capable of bridging these.

**Figure 1.**
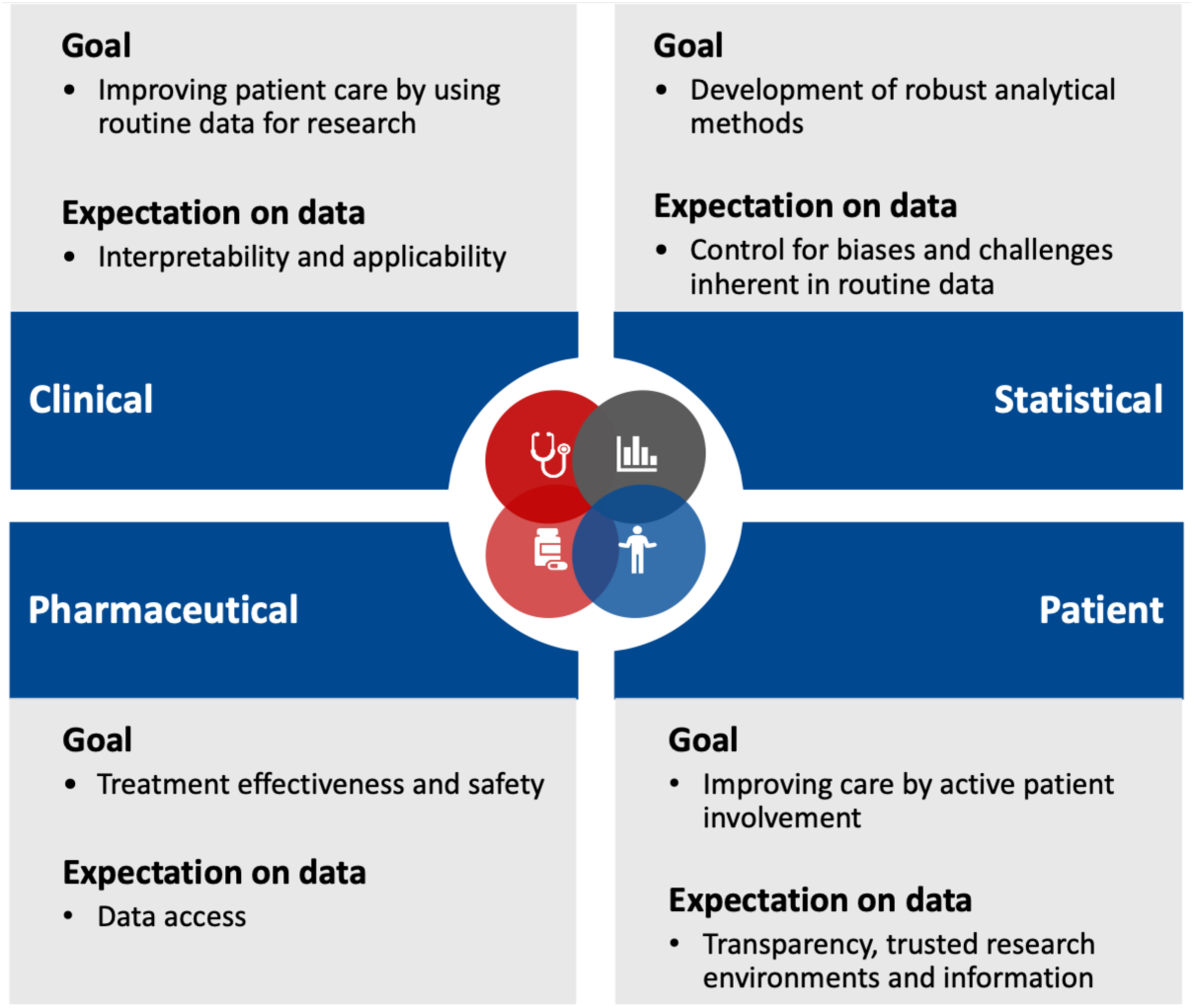
Overview of stakeholder’s perspectives summarizing the goal what should be achieved with routine data and expectations on the data.

### 3.1 Stakeholder perspectives, expectations and needs

#### Clinical perspective: Interpretability and relevance to clinical pathways

From a clinical perspective, the primary value of routine data lies in its potential to inform patient care, based on real world evidence, extend our epidemiological knowledge and to guarantee quality of patient treatment. In addition to hypothesis-driven clinical research, routine data are also a valuable resource for epidemiological surveillance, benchmarking and quality assurance, enabling healthcare providers and policymakers to monitor variations in care and outcomes across institutions and regions. Clinicians require analyses that are interpretable and directly applicable to treatment decisions, ideally aligned with familiar care pathways. For example, stroke registry data in Baden-Württemberg (Germany) have demonstrated that direct admission to comprehensive stroke centers is associated with improved patient outcomes compared with secondary transfers, thereby confirming earlier observational findings and informing referral policies [7]. Similarly, routine data have been used to evaluate off-label intravenous thrombolysis in elderly stroke patients, showing effectiveness in a group excluded from randomized trials [8]. These examples illustrate how analyses rooted in clinical pathways can yield actionable insights. Methodologically, this perspective favors approaches that structure patient trajectories over time, capture intermediate outcomes such as hospital readmission, and allow linkage with patient-reported outcomes to reflect the broader patient journey. Challenges with respect to routine data appear from a statistical point of view when analyzing these data, highlighting the need for close collaboration with statisticians.

#### Pharmaceutical perspective: Regulatory compliance, effectiveness, and safety

For the pharmaceutical sector, the central priority is robust evidence on treatment effectiveness and safety that can complement or substitute randomized controlled trials. Such evidence is particularly relevant when trial results are outdated, infeasible due to ethical or logistical constraints, or impossible in rare or vulnerable populations. Methodologically, the industry perspective requires approaches such as federated learning and target trial emulation, which mimic randomized comparisons in observational data while addressing common sources of bias [9]. Beyond effectiveness, safety endpoints must also be incorporated, for example by representing adverse events as intermediate states within MSMs. These approaches provide evidence that is both relevant for regulatory decision-making and valuable for guiding drug development pipelines. For pharmaceutical industry it becomes challenging to obtain routinely collected data for their studies in adequate quality. In Germany, the so-called “anwendungsbegleitende Datenerhebung” (routine data collection) can be an HTA (Health Technology Assessment) obligation that is issued in case of insufficient data availability for benefit assessment (e.g. in rare diseases) to assess the medication in routine care [10]. Most of these routinely collected data rely on disease registries.

#### Patient perspective: Transparency, PROMs/PREMs, and privacy

Patient representatives emphasized the importance of transparency, trust, and inclusion of patient perspectives in analyses of routine data. Surveys consistently show that many patients are willing to share health data, provided that privacy safeguards are in place, withdrawal from consent is easily possible and potential benefits are clearly communicated [11–14]. However, concerns remain particularly among older individuals, where skepticism is linked to perceived risks of data misuse [13]. In order to increase the willingness to share data, it is essential to communicate both purposes and results of the studies tailored to the target audience. Positive use cases should be used for illustration. Technical and governance solutions, such as trusted research environments, are also required to ensure that data use remains secure and that public trust is maintained [15]. From a methodological standpoint, the integration of patient-reported outcome and experience measures (PROMs and PREMs) alongside clinical data provide patient-orientated research, despite challenges such as incomplete questionnaires or technical barriers. Incorporating PROMs and PREMs, if selection and reporting is clinically relevant [16], enables evaluation of quality of life and satisfaction with care processes, making analyses more relevant from the patient’s perspective.

#### Statistical perspective: Rigor, bias control, and analytical transparency

The statistical perspective centers on ensuring methodological rigor and mitigating the biases inherent in observational routine data. Challenges such as missing information, irregular follow-up, and confounding must be addressed through appropriate analytical strategies. Statisticians emphasize the importance of transparency in assumptions, reproducibility of results, and careful balance between model complexity and interpretability. The statistical role varies across research questions, ranging from primarily consultative role in standard analyses to the application or even development of advanced methods in more complex settings. Time-to-event data are commonly modeled with survival analysis or competing risks methods, but more flexible frameworks are needed to capture sequential trajectories and complex healthcare pathways. MSMs meet these requirements by enabling the estimation of transition probabilities, hazard ratios, and state-specific sojourn times, while maintaining a graphical structure that supports interpretability.

#### Integration across perspectives

The framework illustrates how stakeholder priorities both converge and diverge. Clinicians and patients consistently emphasize privacy, interpretability and transparency, whereas statisticians focus on methodological rigor and the pharmaceutical sector prioritizes regulatory compliance and safety. Despite these different emphases, there is common ground in the need for analytical strategies that capture clinical healthcare pathways, integrate diverse outcomes, and remain transparent in their assumptions. The research question must be framed at the beginning with respect to the primary perspectives of interest (clinical, statistical, pharmaceutical, or patient), which defines the target estimand and informs the choice of an appropriate modeling approach [17,18].

When it comes to modeling event data over time, MSMs stand out as particularly well-suited to operationalize the framework. Their graphical representation of health processes supports clinical interpretability; their flexibility allows the inclusion of intermediate events, adverse outcomes, and patient-reported measures; and their formal statistical structure ensures transparency and rigor. Importantly, they can generate outputs relevant to regulatory contexts while maintaining patient-centered perspectives.

Thus, MSMs provide a unifying methodological exemplar that embodies the principles of the stakeholder-inclusive conceptual framework. In the following section, we illustrate how MSMs can be applied to routine health data to address the priorities of different stakeholder groups.

### 3.2 Multistate models as a methodological bridge across stakeholders

Due to the longitudinal and sequential nature of routinely collected health data, MSMs constitute a suitable modeling approach. A MSM is defined as a finite number of distinct predefined states with transitions between states over the course of time. These states are typically visualized via boxes with possible transitions between states represented by directed arrows. In Figure 2, a prominent example of a MSM, the so-called illness-death-model, with three states is depicted. From the initial health-state a patient can either transit to illness-state or to death-state given the respective transition probabilities. The death-state is absorbing, meaning that no further transitions from that state are possible. The illness-state is referred to as a transient state, as the transitions to another state are feasible. MSMs allow to estimate several outcomes such as overall or progression-free survival accounting for intermittent events, transition probabilities between events, hazard ratios for each transition or even the duration in different states [19]. For more technical details we refer to existing literature [19–21].

**Figure 2.**
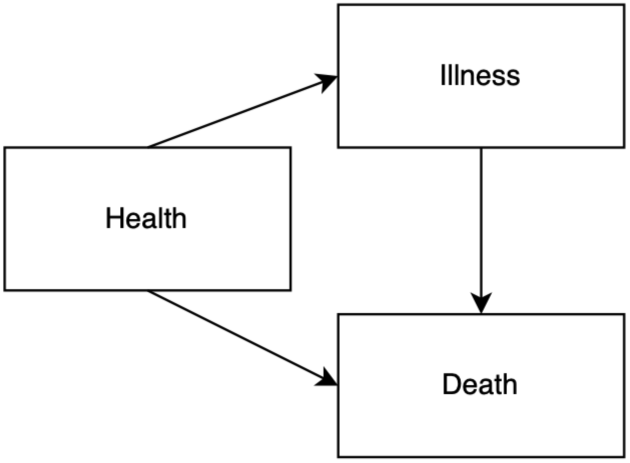
Illustration of an illness-death model, a specific type of multistate model.

Based on the research question of interest and the available data, MSMs can be easily built and adjusted in complexity to appropriately address the research question. A great overview of the flexibility of MSMs and the wide range of questions that can be addressed given the available information in the data is provided by Skourlis et al. [22]. Using Swedish breast cancer registry data including repeated prescriptions on anti-depressants, they progressively refine the MSM, with each more complex version incorporating more information from the available data while providing insights into what the added complexity contributes. In addition, the great flexibility of MSMs is also evident in the versatile applications across various research topics that are relevant to different stakeholders. Specifically, MSMs are suitable to model disease and treatment trajectories, patient’s clinical healthcare pathways over time, or even compare treatments along the clinical care path. To illustrate the interplay of different perspectives, we provide an overview of the different research topics in Additional file 1, supported by example articles. For each article, we outline the research question addressed, the clinical domain and identify the primary perspectives for which the study is most relevant: clinical, pharmaceutical, patient or statistical. Figure 3 illustrates example MSMs with respect to the different research topics while showing the increased complexity and involvement of the statistical perspective. Building on the overview in Additional file 1, the following sections present a summarizing discussion of each research topic.

**Figure 3.**
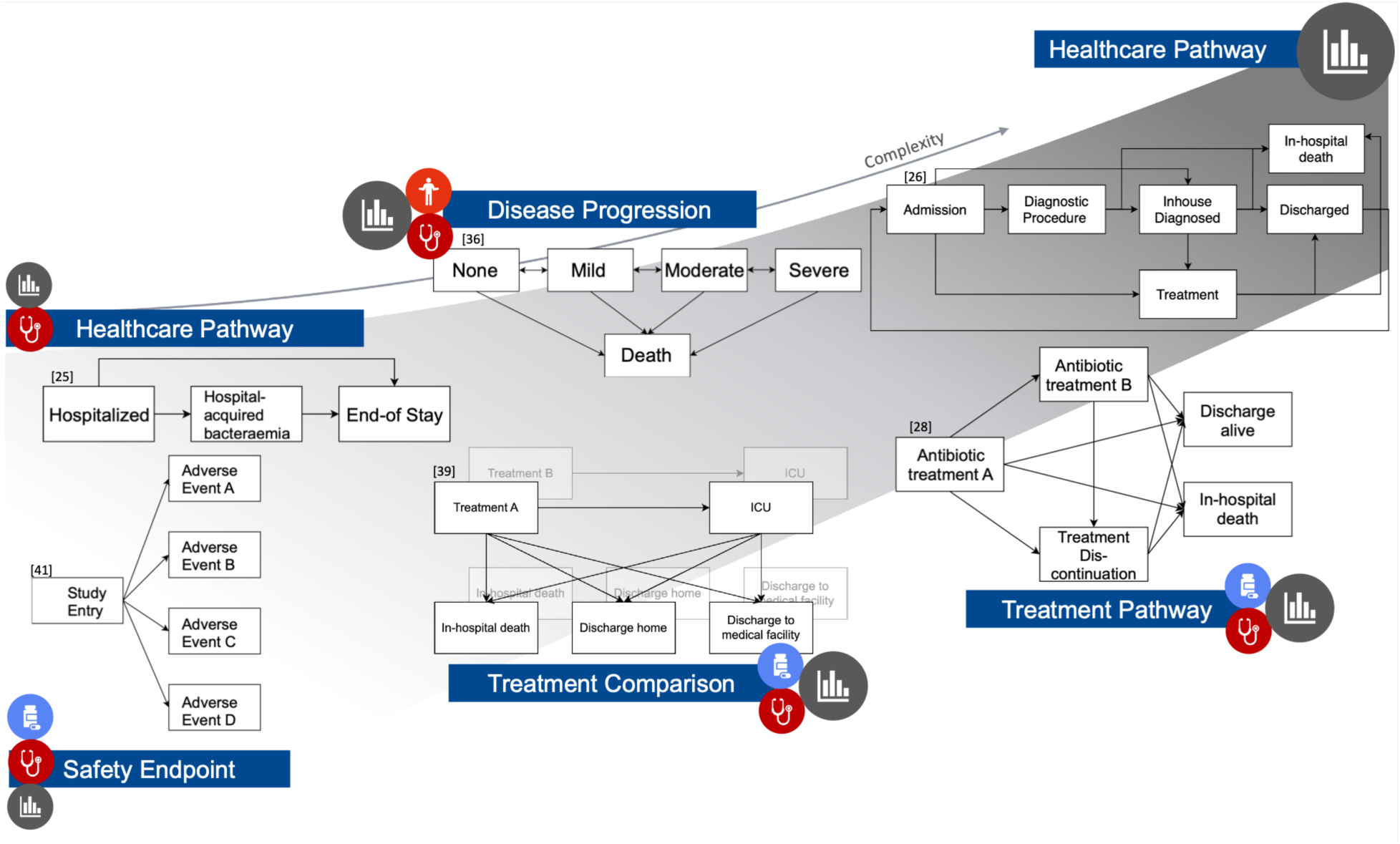
Example multistate models across different research topics, illustrating increasing statistical complexity. The icons next to the research topics, as also used in Figure 1, indicate the primary relevance for each perspective. With increasing complexity of the analysis, the role of the statistician shifts from consultation to methods developments.

#### (i) MSMs for healthcare pathways (clinical, statistical)

A clinical healthcare pathway outlines a patient’s journey in the healthcare system from initial consultation in the clinic to care completion. It encompasses clinical outcomes such as admission, discharge and readmission to the hospital, death, diagnoses as well as procedures and medications to treat a disease. These healthcare pathways allow the investigation of various clinical research questions, for example, regarding length of stay, readmission, or transitions between the ICU and other hospital wards [23–25]. From a statistical perspective, healthcare pathways offer new challenges to solve with recent research dealing with finding similar patient pathways [26,27]. With a simplified pathway of individual care trajectories, multistate modeling approaches can be used to infer hazard rates/ratios for transitioning between healthcare states as well as the duration sojourning in a state. From a clinical point of view, standardizing care processes help decision-making processes and improve efficiency and quality of care. In most cases, however, research questions concentrate on a specific segment of the healthcare pathway. Therefore, the analysis can be reduced to particular components, such as clinical outcomes, treatments, diseases, or their progression.

#### (ii) MSMs for treatment pathways (clinical, pharmaceutical, statistical)

The full healthcare pathway can be further restricted to investigations concerning only the treatment pathway, focusing on the course of a medical treatment or intervention prescribed to a patient with a particular disease. Next to the example of Skourlis et al. [22] mentioned earlier, which is mostly interesting from a clinical perspective, Peng et al. [28] employed a MSM for investigating intermediate events (termination or initiation of a treatment) on clinical endpoints (discharge, death) in critical ill patients under antibiotic treatment. This pharmacometric MSM provided insights into risk factors influencing transitions between clinical states, offering valuable information for patient clinical care. Additionally, this approach contributed to understanding medication dosing and supports further drug development, making it relevant from a pharmaceutical perspective as well.

#### (iii) MSMs for multiple diseases and disease progression (clinical, patient, statistical)

A further narrowing of healthcare pathways may focus solely on disease-state transitions. In this context, the states of a MSM can represent different diseases, for example in a competing risk setting [29,30]. The aim of such studies often is to identify risk/prognostic factors associated with the development of specific diseases making it particularly useful for understanding the etiology and timing of a disease onset for risk stratification and early intervention.

Alternatively, the modeling focus may be placed on the course of a single disease, with states such as progression, complications, response to treatment or death [31–33], rather than distinct disease entities [29,30]. Disease-state trajectories may also be formulated in terms of severity stages [34–38]. In this scenario, analyzing the probability of disease progression and identifying prognostic factors for progression is of greatest interest. From a pharmaceutical perspective, examining disease progression in combination with the treatment course, such as treatment initiation or discontinuation, can also provide valuable insights drug efficacy and potential need for additional treatment.

In the context of disease progression modeling, MSMs are also suitable to actively incorporate patient’s contribution and perspective by the use of survey data or PREMs/PROMS [29,34,36]. By analyzing transition probabilities and mean sojourn times, patients with rapidly worsening symptom perception can be identified, highlighting those in need of targeted clinical intervention.

#### (iv) MSMs for treatment comparison (clinical, pharmaceutical, statistical)

MSMs can also be applied for comparing treatments in complex setting, not only by evaluating treatment effects on final outcomes such as survival or progression-free survival, but also by examining the effects along the pathways involving intermediate events and competing risks [39,40]. This can be applied, for example, to a clinical pathway where ICU stay is modeled as the intermediate event, and different discharge reasons act as competing events alongside death to compare different medications [39]. Such analyses are particularly relevant for treatment decisions and patient management in clinical practice. From a statistical methodological point of view, treatment comparison using observational data is an active area of research aimed at reducing bias through causal inference frameworks, such as target trial emulation.

Another approach to use MSMs for treatment comparison is to examine treatment effects along multiple pathways of the disease process, such as treatment response, disease progression or relapse, and death [31]. This is crucial in oncology research, where treatment effects are still often assessed solely based on overall survival. However, the use of MSMs allows for a more detailed investigation of the disease course and survival by incorporating these intermediate events. Hence, this application is especially valuable from both pharmaceutical and clinical research perspectives.

### (v) MSMs for safety endpoints (pharmaceutical, patient, statistical)

In addition to comparing treatment effects on clinical outcomes or progression, MSMs can be used to analyze the safety of medical interventions by focusing on undesired outcomes, such as complications – commonly referred to as adverse events. These events can be represented as separate states within a MSM. For instance, to assess the effect of treatment discontinuation on several competing pregnancy outcomes specifically relevant for clinical research [41]. Building on MSMs for disease progression, adverse events can also be incorporated as additional states within the model to fully represent the course of the disease. They are not only relevant from a pharmaceutical perspective, but also from a methodological standpoint, as MSMs can help to better understand drug development scenarios [42] by dynamically modeling the disease and treatment effects, providing a rigorous framework for simulation, design evaluation and causal insights. This is particularly interesting for chronic diseases with disease course that are not yet fully characterized [42].

## 4. Discussion

In this work, we present a conceptual framework for modeling routine data that enable the integration of the diverse needs of different stakeholder perspectives. This was accomplished by an interdisciplinary workshop in combination with a targeted literature search, discussing the potential utilization of routine data and identifying MSMs as a potential analytical method to capture the different requirements.

Different stakeholders of the healthcare system often operate from different positions with different goals, expectations and limitations regarding routinely collected data. While the clinical perspective focuses on identifying interpretable and applicable discoveries for clinical care, pharmaceutical industry profits from using routinely collected data to provide evidence on treatment effectiveness and safety to complement or to substitute controlled trials when not feasible. In contrast, the statistical perspective lays focus on methodological development, bias control and challenges inherent in routine data. The willingness to share data is present from patient’s perspective, however, data protection and benefits need to be educated in an easy and understandable way. Although PROMs/PREMs do not count as routinely collected data, the linkage to routine data offers the possibility to actively address the patient perspectives and there exists already initiatives that aim to integrate PROMs into EHR-infrastructure for research purposes [43].

A crucial aspect prior to any model application is the design of the study and the access to routine data. Regarding the study design, inclusion and exclusion criteria, outcomes as well as hypotheses need to be clearly defined involving statisticians’ input from the very beginning. When it comes to access to routine data, clinical institutions, for example, hold routinely collected data and use it for their research purposes. Improving data quality and sharing such data across institutions combined with similarly diligence along the data processing and modeling steps will benefit both patient-centered clinical research and patient care. To proactively meet potential reservations against such complex collaboration approaches, the need for transparency and patient-centered communication are even more important than in research conducted by a single person who also obtained the patient’s consent for participation. It should be emphasized that the data is used for patient-oriented research, involves ethics approval and collaborative processes between the involved stakeholders to ensure clinically relevant analyses.

Even though routine data collection does not require any additional effort, as the collection is already done for patient care and subsequent reimbursement, processing the data comes with extra effort as a lot of data cleaning, plausibility and quality checks are necessary before the actual analysis can proceed. Similar like in registries, data managers for routine data could help overcoming this issue. Yet, PROMs and PREMs are related to additional effort for patients and its acceptance by patients is ambivalent. While a positive attitude towards PROMs is present because patient’s impressions are taken into account [14], the effort required from patients must be justified by a clear rational for data collection [44].

Nevertheless, a central synergy among stakeholders is the collective focus on knowledge generation for improving patient care. With MSMs, different perspectives can be addressed with its flexibility to cover a broad area of research topics and multiple research questions depending on the available data. From the targeted literature search alone, we could already observe that these models had being widely applied to many clinical domains beyond oncology, and to data sources other than trials. We identified selected illustrative examples demonstrating their long-standing and extensive use across diverse research topics and clinical domains, which we summarize in this study. However, routine data come along with limitations as well, also with respect to multistate modeling approaches. Particularly for EHR or administrative data, needed variables may not be documented to adequately address the research question or their inherent characteristics may introduce bias or confounding [32,35,45]. Additionally, the unavailability of detailed event times, e.g. for diagnoses or medication intake, leads to interval censoring, whereas registries and controlled trials usually provide exact timing information [32,35]. However, as MSMs usually need a large sample size also depending on the complexity of the model, routine data appear to be suitable as loads of data are collected during routine care or they can even be linked to other data sources [35].

This study is also subject to limitations: since the workshop took place on a German conference with participants most likely involved in German healthcare system, the priorities, challenges and expectations regarding the use of routine data might be German specific. However, with the support of the targeted literature review, this study is also applicable for other countries. In addition, we did not perform a systematic search because we intended to illustrate the use routine data in MSMs for different research topics and stakeholder perspectives by means of examples. We focused only on a subset of stakeholders of the healthcare system, not taking into account for example the perspectives of outpatient care providers, regulators or payers. First, our motivation comes from the readily available German-wide EHR data from clinical care. This availability is the result of the German Medical Informatic Initiative (MII), which established a standardized infrastructure for the secondary use of routine clinical data and is continued within the Network of University Medicine (NUM) [46]. In contrast, the nationwide usage of outpatient data depends on establishing Germany’s universal electronic health record (opt-out model; ePA) and making it broadly ready and available for research [3,47]. Second, our focus was on clinical decision-making and pharmaceutical implementation, so that including also regulators’ or payers’ perspectives was beyond our scope. Finally, while we acknowledge that other statistical or machine learning methods may be used depending on the research question, our focus was on MSM due to its flexibility and capability in analyzing dynamic longitudinal event data.

## 5. Conclusions

The effective use of routinely collected health data holds great potential to advance both clinical care and pharmaceutical research. Realizing this potential requires not only addressing methodological challenges such as biases, but also acknowledging and integrating the diverse perspectives of stakeholders. MSMs offer a flexible and dynamic framework to investigate different research topics serving as an integrative tool to align diverse perspectives.

## Data Availability

Not applicable.

## Ethics approval and consent to participate

Not applicable

## Consent for publication

Not applicable

## Availability of data and materials

Not applicable

## Authors’ contributions

NB, HB and MP developed the study and workshop concept and design. NB, AD, DK and MW contributed with short keynote speech at the workshop for further discussion. MP wrote the first draft of the manuscript; NB supervised this process. All authors provided additional intellectual content, read and approved the final manuscript.

## Acknowledgements

The authors thank the workshop participants for their lively discussion and participation.

## Supplementary Material

Additional file 1. Overview of example articles

